# The impact of COVID-19 on primary health care and antibiotic prescribing in rural China: qualitative study

**DOI:** 10.1101/2021.03.01.21252602

**Authors:** Tingting Zhang, Xingrong Shen, Rong Liu, Linhai Zhao, Debin Wang, Helen Lambert, Christie Cabral

**Affiliations:** Population Health Sciences, Bristol Medical School, University of Bristol, Bristol, UK; School of Health Services Management, Anhui Medical University, Hefei, China; Centre for Academic Primary Care, Population Health Sciences, Bristol Medical School, University of Bristol, Bristol, UK

**Author notes:** Corresponding author: Dr Tingting Zhang, Population Health Science, Bristol Medical School, University of Bristol, Bristol, UK.

**Keywords:** COVID-19, antibiotic treatment, township health centre, village clinic, epidemic prevention and control

## Abstract

**Introduction:** Primary health care (PHC) system is designated to be responsible for epidemic control and prevention during the outbreak of COVID-19 in China, while COVID-19 suspected cases in PHC are required to be transferred to specialist fever clinics at higher level hospitals. This study aims to understand to impact of COVID-19 on PHC delivery and antibiotic prescribing at community level in the rural areas of central China.

**Methods:** Qualitative semi-structured interviews were conducted with 18 PHC practitioners and seven patients recruited from two township health centres (THCs) and nine village clinics (VCs) in two rural residential areas of Anhui province. Interviews were transcribed verbatim and thematically analysed.

**Results:** Practitioners’ and patients’ views and perspectives on COVID-19 impacts on PHC services and antibiotic prescribing are organised into four broad themes: switch from PHC to epidemic prevention and control, concerns and challenges faced by those delivering PHC, diminished PHC, and COVID-19 as a different class of illness.

**Conclusion:** The COVID-19 epidemic has had a considerable impact on the roles of rural PHC clinics in China that shifted to public health from principal medical, and highlighted the difficulties in rural PHC including inadequately trained practitioners, additional work and financial pressure, particularly in VCs. Antibiotic prescribing practices for non-COVID-19 respiratory tract infections remained unchanged since the knowledge of COVID-19 was not seen as relevant to practitioners’ antibiotic treatment practices, although overall rates were reduced because fewer patients were attending rural PHC clinics. Since COVID-19 epidemic control work has been designated as a long-term task in China, rural PHC clinics now face the challenge of how to balance their principal clinical and public health roles and, in the case of the VCs, remain financially viable.

## INTRODUCTION

The aggressive COVID-19 epidemic has stressed health systems around the world. Primary health care (PHC) systems in countries like the UK and Australia have rapidly responded to COVID-19, including key measures like shifting to remote consultations and providing online training and guidance, whilst facing challenges to manage increased workload and to practice good infection control.[1, 2] In China, PHC practitioners were designated to conduct COVID-19 screening and referral, monitoring, education and publicity by the National Health Commission.[3] The regulations and guidance issued by state government strictly prohibit any consultation or treatment for patients with fever or cough in PHC and require that all identified COVID-19 suspected patients be transferred to specialist fever clinics at designated hospitals.[4] Evidence from a nationwide study showed that over 90% of PHC institutions have enacted these new tasks.[5] Local case studies also demonstrated the key role of PHC practitioners in monitoring suspected COVID-19 cases at community level,[6, 7] including identifying migrant workers returned from high-risk areas and managing their home quarantine over the Spring Festival.[8]

PHC is a relatively weak, but rapidly growing, sector of China’s health care system that provides general medical services and some basic public health services to residents at community level.[9] There are widespread gaps in quality of PHC, particularly in rural areas.[5] Township health centres (THCs) and village clinics (VCs) are the PHC institutions of rural China’s health care system. These frontline health institutions are most rural residents’ first choice when seeking outpatient care;[9] in rural areas, 64.4% of patients chose to visit VCs and 22.4% visit THCs for general illness.[10] Most VCs are independent entities that are non-government owned and financially semi-autonomous, charging fees that are either paid by patients or reclaimed from insurance.[9, 11] The THCs deliver technical guidance and have oversight of the public health work conducted by VCs,[12] but VCs can diagnose and treat patients attending for general illness such as infections. Notably, China’s ‘primary health care’ system differs in important ways from that in high-income settings; VCs offer only outpatient care and are staffed by ‘village doctors’, most of whom do not hold a medical degree but have some medical training, whereas THCs have inpatient wards and a range of outpatient clinics staffed by fully qualified physicians.

The average proportion of antibiotics used for outpatient encounters in PHC institutions was over 50%,[13] which is much higher than the percentage use of antibiotics recommended by the WHO (≤ 30%).[14] In clinical consultations, infection (g*anran*) is usually described as ‘inflammation’ (*yan*), a term that carries connotations of the redness and heat often seen with infections, and which is more readily understood by patients. Antibiotics are commonly called ‘anti-inflammatory medicine’ (*xiaoyan yao*) by practitioners and patients, and frequently prescribed for infectious illness or ‘inflammation’ (*yan*). One study conducted in rural areas of China indicated that, in THCs and VCs, nearly 90% of patients with a respiratory tract infection (RTI) or urinary tract infection (UTI) were prescribed antibiotics.[11]

With the outbreak of COVID-19 and new epidemic control regulations and measures introduced to PHC, we conducted qualitative research on THCs and VCs to shed fresh light on their current situation under COVID-19 epidemic. This study aims to understand to impact of COVID-19 on PHC delivery and antibiotic prescribing at community level in the rural areas of central China.

## METHODS

### Study area and recruitment methods

This study was carried out in two rural residential areas of Anhui province, one in Huangshan municipality and one in Fuyang municipality. Anhui province is located in the northeast of central China and more than half of Anhui population lives in rural areas.[15] Since the beginning of March, the rate of new COVID-19 positive cases in Anhui province has remained stable; as of the 25^th^ November 2020, there have been 992 positive cases. Fuyang has the 3^rd^ highest incidence of all 16 municipalities of Anhui province, with 156 positive cases. Huangshan is a low incidence area in Anhui and only has 9 positive cases confirmed.[16]

PHC practitioners that had participated in the codesign of an intervention for an ongoing trial were invited to participate in this study.[17] Participants were purposively recruited to capture a range of situations, including township and village, larger and smaller clinics, Traditional Chinese Medicine (TCM) and Western Medicine practitioners, and different geographical and COVID-19 incidence areas in Anhui province. Access to participants was secured via senior-level staff at the two selected THCs, who were identified through the existing networks of the research team. Practitioners were contacted or introduced by the senior staff and all agreed to interview. The practitioners then identified a small number of patients, all of whom agreed to be interviewed. In Chinese society with a strong culture of conformity, anonymous and formal approaches to study recruitment are usually associated with suspicion and likely to cause people to either refuse to participate or to provide a socially acceptable account, while trust is engendered by contacts made through personal relationships.[18–20] Our recruitment method was therefore adapted to the cultural context of rural China to avoid low response rates and social desirability biases.[19, 21] Practitioners and patients were recruited from one THC and five VCs in Huangshan municipality, and one THC and four VCs in Fuyang municipality.

### Data collection

Semi-structured interviews in Mandarin were conducted by a bilingual researcher (TZ). Participants were first contacted by the researcher to confirm the interview time. Interviews were then conducted by phone or WeChat, in line with participant preference, and recorded using an encrypted audio-recorder. Interviews were conducted between 15^th^ July and 2^nd^ October 2020. The interviews ranged in length from 14 to 45 minutes.

The semi-structured interviews were guided by topic guides, one for heads of PHC institutions, one for practitioners, and one for patients (See supplemental files). The interviews explored views and experiences of the impact of COVID-19 on PHC delivery. The topic guides covered: impacts on PHC delivery and health care seeking practices; experiences of and understandings of COVID-19; and how the COVID-19 pandemic had affected the use of antibiotics.

### Data analysis

All interviews were transcribed verbatim and thematically analysed in NVivo 12 Pro. A sub-sample of transcripts that captured the differences in participants’ views and experiences were translated into English. Two researchers (TZ & CC) independently read these bilingual transcripts and assigned initial codes to the data; views and practices that recurred within and across participants were allocated into conceptual categories and themes. Researchers then worked collaboratively to develop the initial coding framework. The remaining transcripts were indexed by TZ using the initial coding framework and refinements were made as necessary. Practitioner and patient interviews were analysed separately. Common codes and categories emerged across transcripts of participants and developed into the major themes.

### Ethics statement

This study has been reviewed and approved by the Ethics Committee of Anhui Medical University (reference number 2020H0110). Audio recorded verbal (rather than written) consent was obtained from all participants.

## RESULTS

Our sample consisted of 25 participants, including two township heads and seven township practitioners from two THCs, nine village practitioners, one from each of the nine VCs, together with seven patients from the two THCs. Among 18 practitioners, most (11) were general practitioners, two were TCM practitioners, two were internal medicine physicians, two were heads of THCs and one was practitioner in outpatient department of the THC. Table 1 presents participant characteristics.

**Table 1.** Participant characteristics.

|  |  | Practitioner participants | Patient participants |
| --- | --- | --- | --- |
| **Total number** |  | 18 | 7 |
| **Gender** | Female | 2 | 5 |
|  | Male | 16 | 2 |
| **Age** | 30-39 | 4 | 3 |
|  | 40-49 | 6 | 1 |
|  | 50-60 | 8 | 3 |
| **Education** | No school / Illiterate | 0 | 1 |
|  | Primary School | 0 | 2 |
|  | Secondary School | 0 | 2 |
|  | Technical high school | 9 | 1 |
|  | College | 4 | 1 |
|  | Undergraduate | 5 | 0 |
| **Place of work/residence** | Township | 9 | 1 |
|  | Village | 9 | 6 |
| **Location** | Xiuning County, Huangshan Municipality | 9 | 2 |
|  | Yingquan District*, Fuyang Municipality | 9 | 5 |

\*Participants were from the rural areas of this district

The results representing practitioners’ and patients’ views and perspectives on COVID-19 impacts on PHC services and the use of antibiotics are organised into four broad themes, (i) switch from PHC to epidemic prevention and control, (ii) concerns and challenges faced by those delivering PHC, (iii) diminished PHC, and iv) COVID-19 as a different class of illness.

### Switch from primary health care to epidemic prevention and control

Practitioners reported a major change in their work to focus on COVID-19 control and prevention and away from seeing and treating patients. Clinical services were largely suspended at the beginning of the outbreak and practitioners found the majority of their workload comprised tracing and screening people at high risk for COVID-19 who ‘*returned from Wuhan or returned from Hubei*’ [P14, township internal medicine physician].

‘*During the epidemic prevention and control period, including pharmacies, telephone follow-up, and the tracing and follow-up of fever patients, there were more things to do and they become more complicated and fragmental. Other workload reduced, the main work is related to epidemic prevention and control*.’ [P1, head of THC]

Practitioners, particularly those working in VCs, described being allocated the additional work of reporting and tracing patients with fever identified in their clinics and delivering COVID-19 prevention information to residents in their area: *‘we also need to ensure the prevention and publicity work for local population*.’ [P10, village practitioner]

Before the pandemic, THCs and VCs would diagnose and treat many patients presenting with respiratory infections and fever but after the introduction of epidemic control regulation, they were only allowed to screen and refer patients with suspected symptoms of COVID-19. Practitioners reported a large decrease in patients, particularly in VCs, attributed mainly to the diversion of patients with fever to specialised COVID-19 clinics. One participant said that ‘*the patient amount has reduced two-thirds*’ [P7, township practitioner] in his clinic.

‘*… patient numbers [have] decreased because the authority required that* [we] *cannot see* [patients with] *fever*.’ [P5, village TCM practitioner]

Practitioners had little knowledge and experience of COVID-19 management and felt that information about this was not relevant to them.

‘*The treatment is none of our business, the physical examination is also none of our business. We only need to know that transferring patients immediately if there is certain symptom*.’ [P2, village practitioner]

‘*We’ve not been trained about the treatment* [of COVID-19]. *We mainly focused on preventions and infection control, because* [patients who may need to receive] *treatment were all transferred. This is our focus; we mainly highlighted the awareness of control and prevention*.*’* [P13, head of THC]

### Concerns and challenges faced by those delivering primary health care

Practitioners were very worried about catching the virus at the beginning of the outbreak because they were working on the frontline and initially lacked personal protective equipment (PPE). They faced uncertainty about their exposure from patients in the community: ‘*you contacted too many people at that time and had no idea at all about who may carry the virus*’ [P17, township TCM practitioner]. Some practitioners worried about transmitting the virus to their family and chose to live at their clinics for a time.

‘*Such worry is absolutely there. When this disease first outbreak, we didn’t even have a mask, but we had to go* [to the frontline] *when we were told. We needed to check body temperature for people who returned from other places. Once there is a confirmed case, we will also have to be isolated. How far can you be away from them when checking the temperature? At the beginning, the winter, the supplies are scarce, and the masks are limited*.’ [P9, village practitioner]

‘*Well, I lived at the clinic in that half month and never went back home, since those* [people he checked body temperature] *are all returned from Wuhan, I definitely cannot go back home. … Yes, it is* [for the sake of protecting family], *even we don’t care, we should protect our family, right?*’ [P4, village practitioner]

Practitioners described how COVID-19 and the related changes in regulations had made their work more difficult and stressful. Despite the decrease in patients, their workload had actually increased due to COVID-19 control and prevention work. PPE was initially lacking and then difficult to use: ‘[you] *cannot walk around* [in protective gown] *and* [it] *will be very difficult even to use the toilet*’ [P16, township internal medicine physician].

‘*That’s true that patient numbers decreased, but this thing [the epidemic made you] feel alarmed, so the burden from mental side surely increased*.’ [P9, village practitioner].

‘*Workload should increase, we definitely have a high workload due to COVID-19. To what extent? We need to work day and night to, first, follow-up, and another, check body temperature, of returned people* [from high risk areas], *the more returning people the higher your workload would be*.’ [P10, village practitioner]

One participant felt the work of reporting and transferring all patients with possible COVID-19 symptoms was ‘*not achievable*’ and instead he just asked patients with fever to leave his clinic: ‘*No referral, which will bring a lot of troubles. Just ask him* [patient with fever] *to leave. … Yes, there is a complicated set of procedures for referral*.’ [P5, village TCM practitioner].

VCs experienced financial pressures as patient numbers and fees reduced and they became more reliant on income from the state for provision of public health infection prevention and control services. VCs had to scrap expired medicines purchased in anticipation of the normal level of need, but no financial compensation was available. Practitioners believed that they could maintain their business under such pressure, although it was more difficult than before.

‘*There must be* [impacts]. *Since the patient number in our clinic decreased, it definitely has impact on us*.’ [P3, village practitioner].

‘*These types of medicine were bought with our own money, so will they* [authorities] *care about the expiration? They will not care about it at all. … There is no single penny from subsidy at all, not to mention others. There is nothing, just as same as usual*.’ [P5, village TCM practitioner]

‘*Patients* [number] *did decrease*, [but] *we also worked for public health education and the state ensured the money for this, so our life is guaranteed*.’ [P11, village practitioner]

### Diminished primary health care

VCs were no longer able to treat patients with common infections after the introduction of epidemic control regulation. There was no substitution of face to face with remote consultations (by phone or communications app), which practitioners attributed to the fact that most of their patients were elderly and were not used to remote consultations and/or don’t know how to use smartphones.

‘*As for fever among local people, you know, the kind of cold* [that can easily cause fever] *especially for children. We have more child patients here, and in fact, it is very normal for them to have a fever caused by, for example, respiratory tract infections or tonsillitis, right?*’ [P8, township practitioner]

‘*There is no consultation* [through WeChat or phone]. *Almost all patients here are old people; remote consultation is rare, most of them will directly visit our village clinic*.’ [P12, village practitioner]

Practitioners also reported reductions in antibiotic intravenous drip treatment because of concerns about increased risk of catching COVID-19 when spending time and/or gathering at health institutions: ‘*patients are unwilling to be delayed in township hospitals*’ [P1, head of THC] and ‘*intravenous drips at clinics were not allowed because it would lead patients to gather together, so intravenous drips were stopped at that time*.*’* [P15, township practitioner].

COVID-19 specialist clinics were established in some township hospitals at the start of the epidemic and ‘*most staff were transferred to work for pre-diagnosis and fever clinic’* [P13, head of THC]. However, these regional clinics soon closed to COVID-19 patents since they lacked the qualified staff and equipment required. Patients with respiratory symptoms or fever then had travel even further for treatment.

‘*It was set up at the beginning; however, when the authorities came for inspection, they found these fever clinics did not meet the standards and also not have enough staff. These fever clinics were then withdrawn with only the referral counter remaining*.’ [P1, head of THC]

Patients found it more difficult to access health care as a result of the pandemic. Patients were afraid of being infected with COVID-19 and most patients said they would avoid places they viewed as having a high transmission risk, like health institutions, as a precaution. Patients said they were no longer able to access treatment at township or village level and some said that it was even difficult to get to pharmacies to purchase medicine.

‘*If there’s not a necessary thing, I definitely don’t want to go to hospital. After all there are many patients. What if I was all right but then be infected just because I visited there, right?*’ [P19, technical high school, village, 50 years]

‘*Our township* [level health institutions] *did not treat* [patients with fever], *they even did not prescribe medicine to them. Our local* [health institutions] *did not take any examination on them; they were all transferred to Fuyang City*.’ [P23, illiterate, village, 51 years]

*‘It must be inconvenient to see a doctor, it’s even impossible to go out once I wanted to buy some medicine for my niece*.’ [P22, primary school, village, 46 years]

### COVID-19 as a different class of illness

Practitioners described COVID-19 as different from the RTIs that, pre-pandemic, they had often treated in village and township clinics. This difference was characterised in a range of ways by different practitioners. Many described it as much more serious: ‘*fierce and fatal*’ [P13, head of THC]. Some described it as having a different mode of action on the body or lungs. TCM practitioners understood COVID-19 through plague theory, the TCM theory used to explain illnesses with characteristics of high transmissibility and severity, rather than the ‘hot-natured state’ and ‘hot’ conditions that have been commonly associated with common RTIs: *‘a plague* [Wenyi] *that related to outside environment*’ [P17, township TCM practitioner].

‘*It* [COVID-19] *is also inflammation, but its inflammation came quite fiercely and fast*.’ [P11, village practitioner]

‘*This COVID-19 will quickly cause shock and exhaustion; it’s so severe. If you have general pneumonia, you can recover after admission in the hospital for ten or eight days, but COVID-19 is so severe and cannot be treated by common medicine*.’ [P11, village practitioner]

‘[COVID-19 is] *lung consolidation that is caused by the virus rather than inflammation*’ [P12, village practitioner]

COVID-19 was associated with SARS, also a serious epidemic that was not treated in the routine work of THCs and VCs. Like SARS, COVID-19 was thought to have high transmissibility and severity characteristics that were not commonly associated with normal RTIs and were of particular concern to practitioners and patients. COVID-19 was seen as more severe than SARS, partly due to the more severe and prolonged restrictions imposed to control it.

‘*Like SARS and COVID-19, they all belonged to corona virus, SARS in 2003 also came fiercely and also could lead to death. …Our respiratory tract would also locate many viruses, but generally they do not cause such serious consequences, but this virus* [COVID-19] *is a big issue*.’ [P13, head of THC]

‘[COVID-19 and] *general pneumonia still have certain differences. First of all, for general pneumonia, it is not contagious in general situation. Look at it* [COVID-19], *then it is quite contagious…* [P15, township practitioner]

‘*Yes, this disease is more serious than SARS. SARS can be controlled. See how many days have been spent to control this disease? Isn’t it still there?*’ [P21, college, town, 31 years]

‘*For sure, people will definitely be worried and afraid. In the past, even for SARS period, you didn’t close the road, right? Now the villages are closed, you are not allowed to go out, and then the roads are closed, some places the roads are even cut off so you cannot use them and go out. It feels more serious*.’ [P25, secondary school, village, 35 years]

Practitioners believed that the COVID-19 epidemic had not led to any changes in their antibiotic treatment practices, except that they were seeing lower numbers of patients presenting with infections and therefore the amount of antibiotics prescribed had decreased during the epidemic period. Since practitioners treated COVID-19 as different from the common RTIs seen in their daily work, they did not see their treatment experiences as relevant to COVID-19 or information about COVID-19 treatment as relevant to them.

‘*That is to say, for COVID-19 epidemic’s own impact on the daily use of antibiotics and the consultation, I don’t think it is much different from previous years*.’ [P8, township practitioner]

‘*Once* [the patients] *get fever, the problem is that we cannot prescribe medicine, so antibiotic use surely is less*.’ [P4, village practitioner]

‘*the number of patients dropped sharply and sometimes there were no patients. … Like medicines used in intravenous infusion were likely to be scrapped quite a lot, most of them were antibiotics…*’ [P8, township practitioner]

‘*Village clinics would not be interested in that, doctors in village clinics are impossible to see how antibiotics could be used for COVID-19; they won’t even look at it*’ [P4, village practitioner]

## DISCUSSION

Our study identified a shift of workload and responsibility to epidemic control and prevention in PHC in rural areas of central China over the COVID-19 epidemic period. PHC institutions were not allowed to diagnose or treat any infections with suspected COVID-19 symptoms, and were not familiar with COVID-19 management, which was designated to specialist facilities. Both practitioners and patients reported a decrease in PHC delivered at community level, along with practitioners’ concerns about catching the virus, increased work pressure, and financial pressure particularly in VCs. Regarding the use of antibiotics, practitioners believed that COVID-19 did not change antibiotic treatment practices in PHC, since they had fewer opportunities to treat RTIs and understood COVID-19 as different to common RTIs.

This study shows the key role played by rural PHC practitioners in COVID-19 screening and monitoring. In response to the epidemic, these PHC services considerably increased their public health work (tracing, screening, education), while their provision of general medical services was reduced because patients were referred elsewhere or avoided clinics. This study did not find a shift to remote consulting that has been observed in PHC systems in countries such as the UK.[2] A systematic review found that while other countries maintained essential clinical services through transforming to remote consultations, China’s national guidelines lacked responses to this domain.[22] However, since most rural residents are elders,[23] remote consultations would be difficult to operate in rural areas in China since older residents are reluctant or unable to forego the normal face to face consulting format.

The reassignment of PHC practitioners away from patient treatment and into public health tracing, referral and educational roles highlights the difficulty of a PHC workforce with limited medical training. The quality of diagnosis and treatment at rural PHC has been evaluated to be low,[24] with a high proportion of inadequately trained practitioners.[25] The most well-qualified health practitioners tend to be concentrated in large hospitals and urban areas, contributing to inequities between rural and urban areas in terms of access to better qualified health practitioners.[25, 26]

The COVID-19 epidemic brought considerable additional pressures and stress for practitioners, particularly those working in VCs. There were widespread concerns about catching the virus and PPE shortages across the entire health care system in China,[27, 28] as well as among health workers in other countries.[29] Work pressures and fatigue from the high workload related to paperwork and reporting were reported from practitioners elsewhere in China’s PHC system, and it was suggested that additional staff were need for these low-skilled tasks so that practitioners could focus more on diagnosis and treatment.[28] Our study indicated that, compared with THCs, the reduction of patient numbers and fees caused more obvious financial pressures on VCs. This is related to their financially semi-autonomous status that means the clinic’s overheads, operational costs, and the majority of staff salary need to be covered by income generated from seeing and treating patients. These financial considerations have been identified as a critical reason for village doctors to prescribe antibiotics, including intravenous administration which generates more income,[11] since they are linked to the economic survival of practitioner and clinic. However, practitioners participating in this study reported that their VCs were able to maintain their business despite substantial decreases in patients attending and corresponding drop in antibiotic prescriptions, suggesting the possibility that clinics may be economically viable without the overuse of antibiotics.

Practitioners and patients perceived COVID-19 to be a very different type of illness to other RTIs. This was despite the apparent similarity in symptoms with fever and cough being both key indicators of suspected COVID-19 and, pre-pandemic, frequently taken to indicate an infection/inflammation that needed antibiotic treatment.[11] COVID-19 was associated with SARS because of the similarity of the response of the health care system and national control measures. SARS differed from previous infectious diseases in the rapid spread that hit China’s health care system and caused widespread panic.[30, 31] The national regulations and guidelines on specialised referral and treatment process further emphasised COVID-19 as a unique class of illness. The severity of COVID-19, including characteristics of rapid development, high transmissibility and fatality, are in line with TCM plague (*wenyi*) theory, which assigned COVID-19 to the category of plagues with different properties from other illnesses in TCM, including general RTIs.[32,33] In TCM aetiology, the cause of COVID-19 is epidemic miasma (*liqi*), compared with the six excesses (*liuyin*) (including excesses of wind, cold, summer heat, dampness, dryness, fire) that are related to common cold and fever.[34] COVID-19 was understood as an entirely different types of illness from the RTIs the practitioners commonly diagnosed and treated in their clinics and therefore knowledge of COVID-19 was not seen as relevant to their practice. Antibiotic prescribing practices for non-COVID-19 RTI remained unchanged, although overall rates were reduced because fewer patients were attending rural clinics.

Our qualitative study has provided new and detailed evidence of the impact of COVID-19 on PHC system and antibiotic prescribing in rural China. Our study recruited practitioners and patients from both high and low incidence areas and included a diverse range of rural practitioners from THCs and VCs in various specialities. The number of patients participating in this study was small, but their views added an important perspective on the changes to health care services described by practitioners.

## CONCLUSION

The COVID-19 epidemic has had a considerable impact on PHC in rural China. PHC practitioners took on a key public health role of tracing, screening and educating in rural areas, while their role in seeing and treating patients was reduced, since many patients were diverted to specialist COVID-19 clinics. There was no transition to remote consultations, which was partly attributed to the diversion of patients away from the inadequately trained rural practitioners towards more highly trained medical practitioners in urban areas, and partly to high numbers of elderly patient in rural areas who had neither technology nor inclination for remote consultations. The additional work and risk that PHC practitioners faced placed considerable strain on them, particularly those working in the VCs. Since local outbreak and epidemic control work has been designated as a long-term task in China, rural PHC clinics now face the challenge of how to balance their principal clinical and public health roles and, in the case of the VCs, remain financially viable.

## Supporting information

data collection form

data collection form

data collection form

## Data Availability

The authors confirm that the data supporting the findings of this study are available within the article. For any further enquiries about the data please contact the corresponding author.

## ACKNOWLEDGEMENT

Many thanks to MSc students in Anhui Medical University for their transcribing work. We are grateful to all participants who contributed to this study and to senior staff in local PHC who helped with the recruitment. All authors contributed to, and read and approved, the final version of the manuscript. For any enquiries about the data please contact University of Bristol.

