## Supplementary material for "The impact of COVID-19 on primary health care and antibiotic prescribing in rural China: qualitative study": data collection form

### Topic guide for heads of township hospitals or local health authority officers

#### Introduction and explain the nature and purpose of the study

Thank you for agreeing to take part in the interview. Our study aims to explore how Covid-19 affects the township and village clinics in rural Anhui. We are particularly interested in how health professionals and patients understand the impacts of Covid-19 on their views and practices, and what additional modifications may be needed for our interventions to reduce overuse of antibiotics. The information you provide will help us improve our understanding of Covid-19 and our intervention refinement.

#### Confirm consent to take part

I will start by audio recording your consent to take part in this interview – I will turn my audio recorder on now and ask you the consent questions – please answer yes or no.

1. Do you agree to our conversation being audio recorded?
2. Do you know you are free to stop the interview at any point and you ay skip questions you would prefer not to answer?
3. Do you understand that when we write about our research, we may quote what you said but it will be anonymous and not be possible to identify you?
4. Do you understand that we will keep a record of this interview for future research but without anything that could identify you?

#### Township hospital heads & officers topics

1. Can you describe how has Covid-19 affected this township hospital/ healthcare system in your area? (open question to elicit free account, avoid focussing on antibiotics too early; encourage participants to describe the situation over the entire Covid-19 period)

- *Prompt:*
  - *The use of village clinics, township hospitals and the setting up of fever clinics and emergency department? How long do you expect this situation will last for?*
  - *Staffing, training?*
  - *The seeking help process and treatment of general patients, RTI patients and suspected Covid-19 patients (outpatient and inpatient)? The changes in patient number?*
  - *Local quarantine and social distancing rules, any other protection rules such as face mask, PPE, or facilities of hand wash*
  - *Any concerns: individual safety, workload, financial income*
  - *Liaison and cooperation with other departments*
  - *How has these impacts changed during the epidemic?*

2. How would you understand COVID-19?

- *Prompt: Compared with general pneumonia, or SARS?*

- *Prompt: Fever and cough usually indicate inflammation, do you think COVID-19 is a type of inflammation?*
- *Prompt: How would patients think about this? Anti-inflammatory medicines/antibiotics*

3. What information or guideline have you received on Covid-19? Can you describe any changes in health-policy or regulation in response to Covid-19?

- *Prompt:*
    - *Sources*
    - *Under current situation, how can doctors decide to prescribe antibiotics to RTI patients or patients with Covid-19 suspected symptoms (fever or cough)? Do they rely on blood test? Is there any guideline about it?*

4. We've found from recent studies that people who are weakened by Covid-19 are susceptible to secondary bacterial infections. A study of 191 hospital Covid-19 patients in Wuhan recorded a high death rate (50%) of Covid-19 patients with secondary infections.

i) Have you heard any secondary bacterial infections cases in your area? If so, could you please give me some more details?

ii) How doctors treat these cases? Will antibiotics be used in the treatment of these secondary bacterial infections? If yes, what type of antibiotics were used? Is there any guideline about antibiotic use?

iii) How do you think Covid-19 will affect antibiotic prescribing? And how about antibiotic use for RTIs?

- *Prompt:*
    - *Current and long-term impacts?*
    - *Increase or decrease in antibiotics, changes in how antibiotics given (oral, IV), any new/different medications?*

**As may you know, we are designing an intervention to help doctors to reduce overuse of antibiotics. For example, we've designed a training including the latest evidence about antibiotic use for RTIs, Chinese national guidelines for RTI treatment, and some patient communication techniques. However, after the Covid-19 epidemic, we supposed doctors' views towards antibiotic prescribing would be affected.**

5. How do you think the experience of the Covid-19 epidemic will affect our intervention?
6. What kinds of information or guideline do you need, or do you think would be helpful?
7. How can we include it in our intervention to make it more useful for you or doctors?

- It is entirely voluntary to take part in this research. You are free to withdraw at any time within two weeks without providing any reason, and this will not have any impact on your work.
- All information is confidential. No name or personal information will be mentioned when the research is published.

### 卫生院院长及卫健委人员采访问题

#### 介绍及解释此研究

感谢您同意参与此次采访。我们的研究旨在探讨新冠病毒是如何影响安徽农村地区的乡镇卫生院和村卫生室的。我们对医务人员和患者如何理解新冠病毒对他们的观点和行为的影响非常感兴趣，同时我们想了解在疫情过后，我们需要对以前设计的减少抗生素过度使用的干预措施做哪些额外修改。您提供的信息将帮助我们进一步了解新冠病毒和改进干预措施。

#### 确认同意参与采访

首先，我会录音记录您确认同意参与采访 – 现在我将打开录音笔，并询问您一些关于知情同意的问题-请回答是或否。

1. 您有什么问题吗？您同意参与此研究吗？
2. 您同意我们的谈话被录音吗？

### 问题

1. 您能讲述一下新冠病毒对您所在医院/地区医疗系统的影响吗？（开放性问题，避免过早专注于抗生素；鼓励受访者尝试描述整个新冠肺炎期间的情况）
   - 提示：
     - 村卫生室，乡镇卫生院，以及急诊分诊和发热门诊的设置；您认为这种情况还会持续多久？
     - 医疗卫生机构的人员配备，相关培训
     - 一般患者，呼吸道感染患者和疑似新冠患者的求医过程和治疗; 患者人数变化
     - 当地封城隔离措施，社交距离，以及其他保护措施如口罩，个人防护措施，洗手区等
     - 担忧：个人安全，工作量，财政收入
     - 与其他部门的联络与合作
     - 这些影响随着疫情的发展有所改变吗？

2. 如何理解新冠肺炎？
   - 一般肺炎，SARS?
   - 发烧，咳嗽会有炎症炎症，新冠肺是一种炎症吗？
   - 病人怎么认为？消炎药/抗生素

3. 您收到了哪些有关新冠病毒的信息或指南？为了应对新冠，您发现健康政策或相关法规发生了任何变化吗？
   - 提示：
     - 信息或指南的来源是什么？

- 在当前情况下，医生如何决定给呼吸道感染患者或有新冠疑似症状（发烧或咳嗽）的患者开抗生素？他们会依靠验血吗？是否有任何相关指南？

4. 从最近的研究中我们发现，新冠患者很容易发生继发性细菌感染。一项对武汉市 191 例新冠患者的研究还显示新冠患者继发感染患者的死亡率高达 50％。
   I) 您是否听说过您所在地区的任何继发性细菌感染病例？如果是，您能给我举几个例子或更多细节吗？
   II) 医生如何治疗这些病例？抗生素将用于治疗这些继发性细菌感染吗？如果是，会使用哪种类型的抗生素？有相关抗生素使用的指南吗？
   III) 您认为新冠疫情将如何影响抗生素的使用？ 如何影响抗生素在呼吸道感染上的使用？
      - 提示：
         - 当前和长期的影响？
         - 抗生素的增加或减少，抗生素给药方式的改变（口服，静脉注射），任何新药/其他的药物？

**您可能知道，我们正在设计一项干预措施以帮助减少医生过度使用抗生素。例如，我们设计了一项培训，其中包括有关呼吸道感染使用抗生素的最新证据，中国国家呼吸道感染治疗指南，以及一些患者交流技巧。然而在新冠疫情之后，我们认为医生对抗生素使用的看法将受到一定的影响。**

5. 您认为新冠肺炎的相关经历会如何影响我们所设计的干预措施？
6. 您还需要哪方面的信息或指南，或者您认为哪些信息和指南会对您有所帮助？
7. 您认为我们应如何将这些信息或指南添加在我们的干预措施中，以使其对您或医生更有用？

**-END-**

- 参加此研究纯属自愿。您可以在两周之内随时退出，不需要提供任何理由，并且这不会对您的工作产生任何影响。
- 所有信息都是保密的。在研究结果发表时，任何姓名或个人信息都不会被提及。
