## Supplementary material for "The impact of COVID-19 on primary health care and antibiotic prescribing in rural China: qualitative study": data collection form

### Topic guide for health professionals in township and village clinics

#### Introduction and explain the nature and purpose of the study

Thank you for agreeing to take part in the interview. Our study aims to explore how Covid-19 affects the township and village clinics in rural Anhui. We are particularly interested in how health professionals understand the impacts of Covid-19 on their views and practices, and what additional modifications may be needed for our interventions to reduce overuse of antibiotics that we had discussed in another interview in last year. The information you provide will help us improve our understanding of Covid-19 and our intervention refinement.

#### Confirm consent to take part

I will start by audio recording your consent to take part in this interview – I will turn my audio recorder on now and ask you the consent questions – please answer yes or no.

1. Do you agree to our conversation being audio recorded?
2. Do you know you are free to stop the interview at any point and you ay skip questions you would prefer not to answer?
3. Do you understand that when we write about our research, we may quote what you said but it will be anonymous and not be possible to identify you?
4. Do you understand that we will keep a record of this interview for future research but without anything that could identify you?

#### Health professional topics

1. How has Covid-19 affected how you work in your clinic? (open question to elicit free account, avoid focussing on antibiotics too early; encourage participants to describe the situation over the entire Covid-19 period)

1.1 **Village clinic prompt**

probe around changes in practice:

i) Workload and patient interactions:

- *Could you please describe the process of how patients seek help in your clinic under current circumstance? Whether and how did it change from Jan to June?*
- *Do you know when the specialist fever clinics and referral counts were built? What are they?*
- *Can you meet RTI patients at your clinic now? If no, when did it start? How many patients can you meet per week? Can you describe the latest RTI patient visiting your clinic, when?*
- *Can consultation with RTI patients be provided by other ways, such as remote consultation via e.g. telephone or WeChat?*
- *How long do you expect this situation would last for?*

- *Have you met patients with suspected Covid-19? What did you do?*
- *Did you advised patients to stay away from clinics if have Covid-19/RTI symptoms? Do you know where or which clinics can be visited by RTI patient?*
- *What kinds of care can you provide to patients (e.g. general examination, prescriptions)?*

- *Can you still give a drip in clinics for patient with e.g. mild RTI symptoms or other diseases? Can you describe the latest patient you gave a drip?*

ii) Protective practice

- *What kinds of PPE do you use in your clinic?*
- *Social-distancing rules?*
- *Facilities to wash hand, facilities to sterilise the room and equipment in the clinic?*

Probe around concerns:

i) Do you have any concerns about your own safety while working in the clinics?

ii) Financial implications for clinic

- *As we learned from previous study, clinical service, such as RTI consultations, prescriptions and drips, is an important part contributing to the clinic's incomes; under current situation, how do you think the Covid-19 financially affect your clinic? What is the current financial situation of your clinic? Any necessary expenditure? The clinic's main income?*
- *Do you have any financial support from e.g. local authority?*
- *Do you or other clinics ever tried any methods to protect the income of the clinics?*

iii) Do you have any concerns about the workload and time? Are they decreased or increased? Do you need to report and tract patients only by yourself? Have you asked patients with fever to leave directly?

1.2 **Township hospital prompt**

probe around changes in practice:

i) Workload and patient interactions:

- *Could you please describe the process of how patients seek help in your hospital under current circumstance? Whether and how did it change from Jan to June?*
- *Do you know when did the fever clinics and emergency points particularly facilitated for Covid-19 patients start? Do you think there might be further changes (e.g. stop to use after epidemic)? Do you know any other clinics like these?*
- *Do you work at fever clinics or emergency points? Do you also work at your original clinics for general patients or RTI patients? How do you shift your responsibilities between these two parts?*
- *Have you met patients with suspected Covid-19 at the consultation outside of fever clinics or emergency points? What did you do?*

ii) Protective practice

- *What kinds of PPE do you use in your clinic?*
- *Social-distancing rules?*
- *Facilities to wash hand, facilities to sterilise the clinics and equipment in the clinics?*

Probe around concerns:

i) Do you have any concerns about your own safety for working in your original clinic or fever clinic?

ii) Potential financial concerns? What is the current financial situation of your clinic? Any necessary expenditure? The clinic's main income?

iii) Do you have any concerns if you need to shift your responsibilities between your original clinic and fever clinic?
   - *Patient safety*
   - *Time and workload (Are they decreased or increased? Do you need to report and tract patients only by yourself? Have you asked patients with fever to leave directly?)*

2. How would you understand COVID-19?
   - *Prompt: Compared with general pneumonia, or SARS?*
   - *Prompt: Fever and cough usually indicate inflammation, do you think COVID-19 is a type of inflammation?*
   - *Prompt: How would patients think about this? Anti-inflammatory medicines/antibiotics*

3. What information or guideline have you received on Covid-19?
   - *Prompt: Sources: online, training, or WeChat, from official authority or colleagues?*
   - *Prompt: What are your views on these? Any places that are difficult to follow?*
   - *Prompt (maybe for township hospital only): under current situation, how can doctors decide to prescribe antibiotics to RTI patients or patients with Covid-19 suspected symptoms (fever or cough)? Do they rely on blood test? Is there any guideline about it?*

4. We've found from recent studies that people who are weakened by Covid-19 are susceptible to secondary bacterial infections. A study of 191 hospital Covid-19 patients in Wuhan recorded a high death rate (50%) of Covid-19 patients with secondary infections.

   i) Have you heard any secondary bacterial infections cases in your area? If so, could you please give me some more details?

   ii) How doctors treat these cases? Will antibiotics be used in the treatment of these secondary bacterial infections? If yes, what type of antibiotics were used? Is there any guideline about antibiotic use?

   iii) How do you think Covid-19 will affect antibiotic prescribing? And how about antibiotic use for RTIs?
   - *Prompt: current and long-term impacts?*
   - *Prompt: increase or decrease in antibiotics, changes in how antibiotics given (oral, IV), any new/different medications?*

**As you know, we are designing an intervention to help doctors to reduce overuse of antibiotics. For example, we've designed a training including the latest evidence about antibiotic use for RTIs, Chinese national guidelines for RTI treatment, and some patient communication techniques. However, after the Covid-19 epidemic, we supposed doctors' views towards antibiotic prescribing would be affected.**

5. How do you think the experience of the Covid-19 epidemic will affect our intervention?
6. What kinds of information or guideline do you need, or do you think would be helpful?
7. How can we include it in our intervention to make it more useful for you?

- It is entirely voluntary to take part in this research. You are free to withdraw at any time within two weeks without providing any reason, and this will not have any impact on your work.
- All information is confidential. No name or personal information will be mentioned when the research is published.

### 乡镇卫生院及村卫生室医务人员采访问题

#### 介绍及解释此研究

感谢您同意参与此次采访。我们的研究旨在探讨新冠病毒是如何影响安徽农村地区的乡镇卫生院和村卫生室的。我们对医务人员如何理解新冠病毒对他们的观点和行为的影响非常感兴趣，同时我们想了解在疫情过后，我们需要对以前设计的减少抗生素过度使用的干预措施做哪些额外修改，这些措施我们曾在去年的一次采访中与您要论过。您提供的信息将帮助我们进一步了解新冠病毒和改进干预措施。

#### 确认同意参与采访

首先，我会录音记录您确认同意参与采访 – 现在我将打开录音笔，并询问您一些关于知情同意的问题-请回答是或否。

1. 您同意我们的谈话被录音吗？
2. 您知道您随时可以自由地停止采访，并且跳过任何您不想回答的问题吗？
3. 您了解当我们撰写有关研究的文章时，我们可能会引用这次谈话的内容，但会是匿名并且不会包含任何您身份的任何信息吗？
4. 您了解我们会保留这次采访的记录以备将来研究，但不会保留任何可以识别出您身份的信息吗？

## 问题

1. 您能讲述一下新冠病毒对您在医院/诊所的工作的吗？（开放性问题，避免过早专注于抗生素；鼓励受访者尝试描述整个新冠肺炎期间的情况）

1.1 村卫生室提示

探讨日常活动中的变化：

i) 工作量和患者互动：

- 您能否描述一下在当前情况下，患者是如何来您的诊所寻求帮助？从一月到现在，这个过程是否有所改变，如何变化的？
- 您知道专门为新冠肺炎患者提供服务的发热门诊和预诊分诊是什么时候建立的吗？有哪些？
- 呼吸道感染患者现在可以来您的诊所就诊吗？是什么时候开始的？一周大概多少个？上个呼吸道感染的病人是什么时候？什么情况
- 您是否可以通过其他方式向这些患者提供咨询，例如通过打电话或者微信？
- 您预计这种情况会持续多久？

- 您是否曾经遇到过疑似新冠肺炎的患者？您是如何处理的？
- 如果患者出现呼吸道感染/疑似新冠的症状，您是否曾建议患者不要来诊所？你知道需要建议他们去哪里或哪个诊所吗？
- 您可以为患者提供哪些医疗服务（例如常规检查，处方）？

- 您还能在诊所为患者吊水吗？例如轻微呼吸道感染患者或患其他疾病的患者。上次给病人吊水是什么时候， 什么情况？

ii) 防护措施

- 您在诊所中使用哪些个人防护用品？
- 是否与其他人保持一定的距离（社交距离）？
- 洗手设备，对诊所房间和仪器进行消毒的设施？

探讨担忧：

i) 您在诊所工作时是否对自己的个人安全有任何担忧？

ii) 对诊所的财务收入的影响

- 根据我们从前的研究，医疗服务例如为呼吸道感染患者看病，开处方或打点滴，都是诊所收入的重要组成部分；在当前情况下，您认为新冠疫情对您诊所的经济收入有何影响？目前的支出收入情况？哪些必要支出（水，电，人员）？主要收入来源？
- 您是否有来自其他方面的财务支持？比如当地政府？
- 您或其他诊所是否曾尝试过任何方法来确保或支持诊所的收入？

iii) 您是否对时间和工作量有什么担忧？和原来相比增加或是减少？自己上报自己追踪？是否会让发热患者直接离开？

1.2 乡镇卫生院提示

探讨日常活动中的变化：

i) 工作量和患者互动：

- 您能否描述一下在当前情况下，患者是如何在您的医院寻医问药的？从一月到六月，这个过程是否有所改变，如何变化的？
- 您知道专门为新冠肺炎患者提供服务的发热门诊和急诊是什么时候建立的吗？您认为这些部门在将来会有什么变化（例如在疫情结束后停止使用）？您知道其他类似的诊所吗？
- 您是否在发热门诊或急诊工作？您是否同时还在普通门诊接诊一般病人或者呼吸道感染病人？这两部分的工作您是如何安排的？
- 您是否在普通门诊/发热门诊或急诊之外遇到了意思新冠患者？您是如何处理的？

ii) 防护措施

- 您在诊所中使用哪些个人防护用品？
- 是否与其他人保持一定的距离（社交距离）？
- 洗手设备，对诊所房间和仪器进行消毒的设施？

探讨担忧：

iv) 在发热门诊或普通门诊工作时，您是否担心自己的个人安全？

- 潜在财务问题的担忧？目前的支出收入情况？哪些必要支出（水，电，人员）？主要收入来源？

v)　您是否对如何安排同时在普通和发热门诊工作有任何担忧?

- 患者安全
- 时间和工作量(自己上报自己追踪?和原来相比增加或是减少?是否会让发热患者直接离开?)

2. 如何理解新冠肺炎?
   - 一般肺炎，SARS?
   - 发烧，咳嗽会有炎症炎症，新冠肺是一种炎症吗?
   - 病人怎么认为?消炎药/抗生素

3. 您收到了哪些有关新冠病毒的信息或指南?
   - 提示:
     - 信息来源:来自网络，微信，来自官方政府，同事?
     - 对指南有什么看法?哪些地方难以服从?
     - (可能只试用于乡镇卫生院)在当前情况下，医生如何决定给呼吸道感染患者或有新冠疑似症状(发烧或咳嗽)的患者开抗生素?他们会依靠验血吗?是否有任何相关指南?

4. 从最近的研究中我们发现，新冠患者很容易发生继发性细菌感染。一项对武汉市 191 例新冠患者的研究还显示新冠患者继发感染患者的死亡率高达 50%。

I)　您是否听说过您所在地区的任何继发性细菌感染病例?如果是，您能给我举几个例子或更多细节吗?

II)　医生如何治疗这些病例?抗生素将用于治疗这些继发性细菌感染吗?如果是，会使用哪种类型的抗生素?有相关抗生素使用的指南吗?

III)　您认为新冠疫情将如何影响抗生素的使用? 如何影响抗生素在呼吸道感染上的使用?

- 提示:
  - 当前和长期的影响?
  - 抗生素的增加或减少，抗生素给药方式的改变(口服，静脉注射)，任何新药/其他的药物?

**您应该已经知道，我们正在设计一项干预措施以帮助减少医生过度使用抗生素。例如，我们设计了一项培训，其中包括有关呼吸道感染使用抗生素的最新证据，中国国家呼吸道感染治疗指南，以及一些患者交流技巧。然而在新冠疫情之后，我们认为医生对抗生素使用的看法将受到一定的影响。**

5. 您认为新冠肺炎的相关经历会如何影响我们所设计的干预措施?
6. 您还需要哪方面的信息或指南，或者您认为哪些信息和指南会对您有所帮助?
7. 您认为我们应如何将这些信息或指南添加在我们的干预措施中，以使其对您更有用?

**-END-**

- 参加此研究纯属自愿。您可以在两周之内随时退出，不需要提供任何理由，并且这不会对您的工作产生任何影响。
- 所有信息都是保密的。在研究结果发表时，任何姓名或个人信息都不会被提及。
