## Supplementary material for "The impact of COVID-19 on primary health care and antibiotic prescribing in rural China: qualitative study": data collection form

### Topic guides for patients in township hospitals and village clinics

#### Introduction and explain the nature and purpose of the study

Thank you for agreeing to take part in the interview. Our study aims to explore how Covid-19 affects the township and village clinics in rural Anhui. We are particularly interested in how patients understand the impacts of Covid-19 on their views and practices, and what additional modifications may be needed for our leaflet designed to help patients not use antibiotics for common colds and cough. The information you provide will help us improve our understanding of Covid-19 and our intervention refinement.

#### Confirm consent to take part

I will start by audio recording your consent to take part in this interview – I will turn my audio recorder on now and ask you the consent questions – please answer yes or no.

1. Do you agree to our conversation being audio recorded?
2. Do you know you are free to stop the interview at any point and you any skip questions you would prefer not to answer?
3. Do you understand that when we write about our research, we may quote what you said but it will be anonymous and not be possible to identify you?
4. Do you understand that we will keep a record of this interview for future research but without anything that could identify you?

#### Patient Topics

1. Under what circumstances would people suspect that they are COVID-19 infected?
   How about and what did people do if they have some RTI symptoms, such as runny nose, sore throat, cough, and fever?
   - *Will they buy medicine from a pharmacy & if so, describe experience? Any antibiotics?*
   - *Will they seek help from a doctor & if so, describe experience? Village clinic, township health centre, or fever clinic? Why?*

2. How has Covid-19 affected you? (elicit free account)
   - *Probe any experience of suspected Covid-19: what did they do?*
     - *Did they buy medicine from a pharmacy & if so, describe experience? Any antibiotics?*
     - *Did they go to a clinic & if so, describe experience?*
     - *Did they face any barrier to attending a clinic (e.g. told to stay home with Covid-19 symptoms or go to fever clinics)? & if so, what did they do?*
     - *Were they tested and admitted to hospital & if so, describe experience?*
     - *Did they self-isolate? How did this work & were there barriers (food supplies & medicine, work, family or other household members, caring responsibilities, finance, mental wellbeing)*
- *Probe if no experience of suspected Covid-19:*
  - *Experience of local quarantine? Is this compulsory?*

    - *What if people need to see a doctor or get medicine during the local quarantine?*
    - *Did you meet any difficulties? (food supplies & medicine/work/isolating with other family members/caring responsibilities/financial issues/mental health)*
    - *Did you ask help from local authority? What happened?*
- *Probe around wider experiences:*
    - *illness in family or wider social network? Any changes compared with pre-pandemic?*
    - *The current process of visiting a doctor? Would you avoid going outside, gathering, or visiting a doctor? Would you purchase and store medicines from clinics or pharmacies?*
    - *ability to protect self? Would doctor advise anything?*
- *Probe around concerns:*
    - *safety of self and loved ones? Any protective measures?*
    - *impact of restrictions*

3. Can you tell me in your own words what you think Covid-19 is?
    - *Probe around:*
        - *ideas of inflammation / infection,*
        - *typical symptoms,*
        - *severity or threat posed,*
        - *who is susceptible*
        - *compared with SARS*

4. What do you think are the best ways to **prevent** Covid-19? Do you think antibiotics would be helpful?

5. What do you think are the best ways to **treat** Covid-19? Do you think antibiotics would be used for an RTI or Covid-19?

- *If yes, probe around: what kinds of, where obtained from, why they chose this medicine*

6. What information or guideline have you received about Covid-19?

- *Prompt: Sources: online or WeChat, from official authority or colleagues/family & friends? Any about antibiotics use? Its impacts on antibiotics use on you*

**We are designing an intervention to help to reduce overuse of antibiotics. For example, we created a leaflet to help patients understand that they should not use antibiotics for common colds and similar RTI infections. However, after the Covid-19 epidemic, we supposed patients' views towards antibiotic use would be affected.**

7. How do you think the experience of the Covid-19 epidemic will affect your thoughts on the leaflet?

8. What kinds of information or guideline do you need, or do you think would be helpful?

9. How can we include it in our intervention to make it more useful for you?

- It is entirely voluntary to take part in this research. You are free to withdraw at any time within two weeks without providing any reason, and this will not have any impact on your work.
- All information is confidential. No name or personal information will be mentioned when the research is published.

#### 乡镇卫生院及村卫生室患者采访问题

##### 介绍及解释此研究

感谢您同意参与此次采访。我们的研究旨在探讨新冠病毒是如何影响安徽农村地区的乡镇卫生院和村卫生室的。我们对患者如何理解新冠病毒对他们的观点和行为的影响非常感兴趣，同时我们想了解在疫情过后，我们需要对以前设计的一份向患者宣传不要在感冒或咳嗽时使用抗生素/消炎药的传单做哪些额外修改。您提供的信息将帮助我们进一步了解新冠病毒和改进干预措施。

##### 确认同意参与采访

首先，我会录音记录您确认同意参与采访 – 现在我将打开录音笔，并询问您一些关于知情同意的问题，请您回答是或否。

1. 您同意我们的谈话被录音吗？
2. 您知道您随时可以自由地停止采访，并且跳过任何您不想回答的问题吗？
3. 您了解当我们撰写有关研究的文章时，我们可能会引用这次谈话的内容，但会是匿名并且不会包含任何您身份的任何信息吗？
4. 您了解我们会保留这次采访的记录以备将来研究，但不会保留任何可以识别出您身份的信息吗？

##### 患者问题

1. 最近人会在什么情况下会怀疑自己感染了新冠？
   如果人们出现一些呼吸道感染的症状，例如流鼻涕，嗓子疼，咳嗽和发烧，他们会怀疑自己感染了新冠肺炎吗？他们会怎么做？
    - 他们会从药店购买药品吗？如果是，请描述相关经历。有抗生素/消炎药吗？
    - 他们会寻求医生的帮助吗？卫生室、卫生院、发热门诊？为什么?

2. 新冠肺炎对您有何影响？（开放性问题，避免过早专注于抗生素）
   - 探讨疑似新冠肺炎感染的相关经历：他们做了什么？
     - 他们是否从药房购买药品？如果有请讲述相关经历。有买抗生素/消炎药吗？
     - 他们是否去过诊所？如果有请讲述相关经历。
     - 他们在去诊所就诊时是否面临任何阻碍（例如被告知如有新冠症状要留在家里或去发热门诊）？如果有，他们会做什么？
     - 他们是否接受了检测？是否入院？如果有请讲述相关经历。
     - 他们是否进行了自我隔离？如何进行以及遇到了什么困难（食品和药品供应，工作，如何与其他家庭成员隔离，如何照顾家人，经济财务问题，精神健康）
   - 如果没有相关经历：
     - 居家隔离的经历？要求是强制的吗？
     - 居家隔离的时候需要看病或配药会怎么做？

    - 遇到了什么困难（食品和药品供应；工作；如何与其他家庭成员隔离，如何照顾家人；经济财务问题；精神健康）
    - 有跟镇子里提要求吗？结果怎么样？

- 探讨更广泛的经历：
    - 疾病对家庭和社会交往造成的影响；和以前有什么变化？
    - 现在的看病流程是什么样的？会尽量避免出门，避免扎堆，避免看病吗？会去药房或医院买药预防，储备吗？
    - 如何自我保护; 医生会交代什么？
- 探讨担忧：
    - 自己和亲人的安全；采取什么措施？
    - 封城所带来的影响

3. 您能用自己的话告诉我您认为新冠肺炎（新冠病毒）是什么吗？
    - 探讨：
        - 对炎症/感染的看法
        - 典型症状
        - 严重程度，危害
        - 哪些人更加容易感染
        - 和非典相比

4. 您认为**预防**新冠肺炎的最佳方法是什么？您认为使用抗生素/消炎药会有用吗？

5. 您认为**治疗**新冠肺炎的最佳方法是什么？您认为抗生素/消炎药是否可以用在呼吸道感染或新冠肺炎？
    - 如果是，探讨：是什么抗生素/消炎药，从何处获得，为什么选择这种药物？

6. 您收到了哪些有关新冠病毒的信息或指南？
    - 提示：来源：来自网络或微信， 来自官方政府，同事，朋友/家人？
      这些信息或指南有和抗生素/消炎药使用相关的吗？它对您使用抗生素/消炎药有何影响？

**我们正在设计一项干预措施以帮助减少抗生素/消炎药的过度使用。例如，我们已经制作了一份传单，来帮助患者了解普通感冒和类似呼吸道感染不应使用抗生素/消炎药。但是在新冠肺炎流行之后，我们认为患者对抗生素使用的看法将受到一定的影响。**

7. 您认为新冠肺炎的相关经历会如何影响您对我们传单的看法？
8. 您还需要哪方面的信息或指南，或者您认为哪些信息和指南会对您有所帮助？
9. 您认为我们应如何将这些信息或指南添加在我们的干预措施中，以使其对您更有用？

**-END-**

- 参加此研究纯属自愿。您可以在两周之内随时退出，不需要提供任何理由，并且这不会对您的工作产生任何影响。
- 所有信息都是保密的。在研究结果发表时，任何姓名或个人信息都不会被提及。
